# Gut microbiome diversity measures for metabolic conditions: a systematic scoping review

**DOI:** 10.1101/2021.06.25.21259549

**Authors:** Chatpol Samuthpongtorn, Tanawin Nopsopon, Krit Pongpirul

## Abstract

**Objective:** Evidence on the association between the gut microbiome and metabolic conditions has been increasing during the past decades. Unlike the straightforward identification of beneficial non-pathogenic bacteria as a potential probiotic for clinical use, the analysis of gut microbiome diversity is more complex and required a better understanding of various measures. We aim to summarize an elaborated list of gut microbiome diversity measures.

**Design:** Systematic search was conducted in three databases: PubMed, Embase, and Cochrane Central Register of Clinical Trials for randomized controlled trials, quasi-experimental and observational studies for the relationship between gut microbiota and metabolic diseases published in 2019 with the English language.

**Results:** The measurement methods of alpha diversity and beta diversity were explored. Of 5929 potential studies, 47 were included in the systematic review (14632 patients). Of 13 alpha diversity measures, the Shannon index was the most commonly used in 37 studies (78.7%), followed by Chao1 index (19 studies), Operational Taxonomic Unit (OTU) richness (15 studies), Simpson index (13 studies), and Abundance-based Coverage Estimators (ACE) index (10 studies). Of 2 beta diversity measures, the UniFrac was the most commonly used in 24 studies (unweighted 17 studies and weighted 16 studies), followed by Bray-Curtis dissimilarity (16 studies).

**Conclusion:** Various measurement of gut microbiome diversity have been used in the literature. All measurements have unique characteristics, advantages, and disadvantages which lead to different usage frequency. The measures were chosen considering cost, simplicity, and types of research.

**Significance of this study:** *What is already known on this subject?:*

▸ Alpha diversity, including Shannon index diversity, chao1 diversity, etc., is the average species diversity within a habitat type at a local scale while beta-diversity, such as Bray-Curtis dissimilarity and UniFrac, indicates the differentiation between microbial communities from different environments.
▸ Alpha- and beta-diversity are the two most diverse measures of gut microbiota diversity with no consensus on which measurement methods should be used in metabolic condition study.

*What are the new findings?:*

▸ Distinct characteristics, advantages, and disadvantages of each microbiome diversity measurement method lead to a variety of usage frequencies in metabolic condition studies. Shannon diversity is the most widely used alpha diversity while there is no predilection for beta diversity.

*How might it impact on clinical practice in the foreseeable future?:*

▸ Further researchers on metabolic condition with microbiome diversity measurement will have impartial evidence on which measurement methods are most rationally appropriate for their studies regarding simplicity, cost, and efficacy.

## INTRODUCTION

Gut microbiota are microorganisms that live in several areas of the body, especially in the gastrointestinal tract. The number of human microbiota, including bacteria, fungus, and virus is approximately 1,000,000,000 - 10,000,000,000 microbial cells, with the ratio of microbial cells to human cells being 1:1.^1 2^ The dominant bacterial phyla in the human gastrointestinal tract are Firmicutes, Bacteroidetes, Actinobacteria, and Proteobacteria.^3^ Current research has found associations between microbiota and systemic diseases, particularly type I and type II diabetes, obesity, and metabolic syndrome which related with immune response processes.^4^

Nowadays, the identification of dominant microbial communities is increasing with the invention of high throughput sequencing technology. The most important and widely used diversities are alpha-diversity and beta-diversity.^5^ Alpha diversity, including Shannon index diversity, chao1 diversity, etc., is the average species diversity within a habitat type at a local scale while beta-diversity, such as Bray-Curtis dissimilarity and Unifrac, indicates the differentiation between microbial communities from different environments.^6^ Both diversities consider two aspects of a community: the number of different organisms in a sample, and the range of abundances for each one.^7^

Many researchers have found the relationship between gut microbiota and metabolic diseases by diversity analysis.^8^ However, there was no systematic study focused on the most widely used method for diversity measurement of the association between gut microbiota and metabolic diseases. This systematic scoping review aimed to discuss and compare the measurement methods of microbiome diversities that are widely used in current research.

## MATERIALS AND METHODS

### Registration of protocol

This study was conducted following the recommendations of the Preferred Reporting Items of Systematic Reviews and Meta-Analyses Extension for Scoping Review (PRISMA-ScR) statement. We registered the systematic review with OSF The Open Science Framework (registration: osf.io/ux2fs).

### Data sources and searches

PubMed, Embase, and Cochrane Central Register of Clinical Trials were used to search for articles published in 2019 in the English language. The terms “gastrointestinal microbiome”, “gut microbiome”, “microbiota”, and “microflora” were used in combination with “diversity”, “richness”, “evenness”, and “dissimilarity” as the keywords for literature search along with their synonyms. The search strategy is presented in detail in the online supplemental appendix 1. Additionally, the reference lists of included articles were searched, as well as related citations from other journals via Google Scholar.

### Study selection

For this systematic scoping review, we worked with an information specialist to design an appropriate search strategy to identify original peer-reviewed articles of randomized controlled trials, quasi-experimental, and observational studies evaluating gut microbiome diversity in patients with a diagnosis of metabolic disease including metabolic syndrome, diabetes mellitus, hypertension, dyslipidemia, obesity, and nonalcoholic fatty liver disease (NAFLD). Article screening was done by two independent reviewers (CS and TN) for eligible studies. Discrepancies between two reviewers were resolved by consensus.

### Data extraction

Data extraction was done by two independent reviewers (CS and TN) for published summary gut microbiome diversity index. Discrepancies between two reviewers were resolved by consensus. We extracted the following data: (1) study characteristics (authors, study type, journal name, contact information, country, and funding), (2) patients characteristics (sample size, type of metabolic disease, and mean age), (3) outcomes (measurement methods of alpha and beta diversity of gut microbiome) as well as any other relevant information. All relevant text, tables, and figures were examined for data extraction. We contacted the authors of the study with incompletely reported data. If the study authors did not respond within 14 days, we conducted analyses using the available data.

### Data synthesis and analysis

The primary outcome was measurement methods of alpha and beta diversity of gut microbiome. We synthesized the overall usage of gut microbiome diversity index whether alpha and beta diversities had been measured in included studies and which index had been used. We then provided subgroup analyses based on study design, type of metabolic disease, geographical location, and country income.

## RESULTS

### Study selection

The database search identified 5,929 potential records. After removing duplicates, 4,119 titles passed the initial screen, and 535 theme-related abstracts were selected for further full-text articles assessed for eligibility (figure 1). A total of 488 were excluded as the following: 338 non peer-reviewed, 65 wrong outcomes, 25 protocol, 22 wrong publication year, 8 duplicate, 8 in vitro, 7 wrong population, 6 review articles, 4 letter to editor, 3 non-English, and 2 editorial. 47 studies were eligible for the data synthesis.

**Figure 1.**
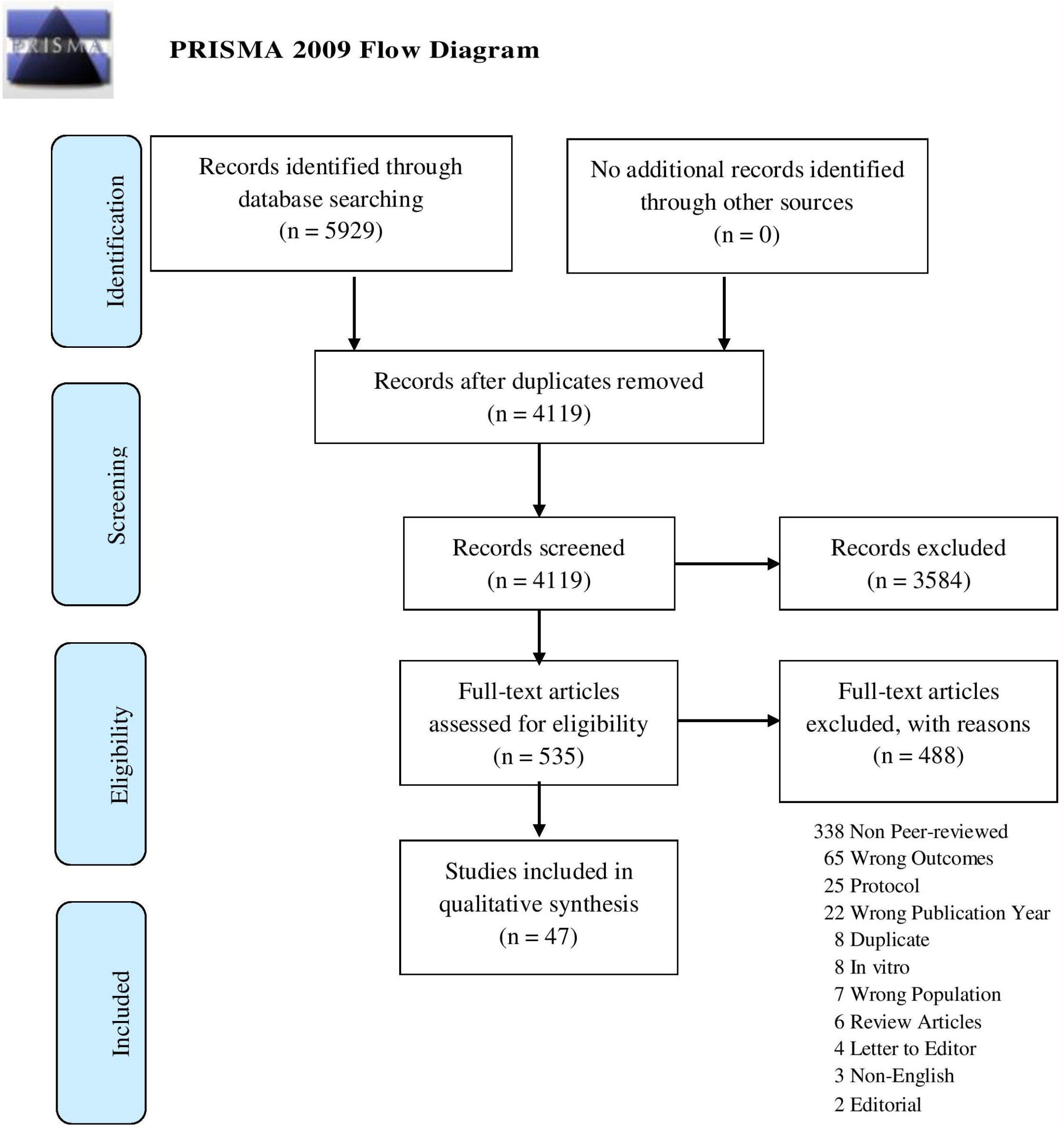
Flow chart diagram presenting the study selection with Preferred Reporting Items for Systematic Reviews and Meta-analyses (PRISMA) guidelines

### Study characteristics

Of 47 included studies, there were 33 observational studies,^9-41^ nine randomized controlled trials,^42-50^ and five quasi-experimental studies (table 1).^51-55^ The number of patients per study ranged from 12 to 6,627, with a total of 14,632 patients. The mean age of patients varied from day of life 3 to 69 years old. There were 28 studies focused on obesity,^3 11 12 16 18 22 24-28 30-34 36 38 39 43 44 46 49-53 55^ 12 studies on type II diabetes mellitus,^3 10 14 15 17 21 26 40 43 45 47 55^ four studies on hypertension,^13 24 37 40^ four studies on NAFLD,^22 33 38 41^ three studies on metabolic syndrome,^31 48 51^ three studies on gestational diabetes mellitus,^19 20 29^ two studies on dyslipidemia,^23 42^ and a study on type I diabetes mellitus.^9^

**Table 1.** Characteristics of Included Studies

| Author | Country | Income Level | WHO Regions | Study design | n | Population | Mean age (SD) |
| --- | --- | --- | --- | --- | --- | --- | --- |
| Robinson<sup>11</sup> | Australia | HICs | Western Pacific | Observational | 22 | 22 Obesity/Overweight | with Ketonuria Med 33 (IQR 29-38), without Med 32 (IQR 29-33) |
| Harbison<sup>9</sup> | Australia | HICs | Western Pacific | Observational | 88 | 47 T2DM, 41 Control | Med 10.9 (Range 4.8-26.7) |
| Horvath<sup>43</sup> | Austria | HICs | European | RCT | 26 | 26 T2DM with Obesity | 60 (NR) |
| Liu T<sup>10</sup> | China | UMICs | Western Pacific | Observational | 6627 | 495 IFG, 304 T2DM, 5828 Control | 51.8 (NR) |
| Lv<sup>12</sup> | China | UMICs | Western Pacific | Observational | 28 | 9 Overweight/Obesity, 10 Normal, 9 Lean | 22.03 (1.32) |
| Mushtaq<sup>13</sup> | China | UMICs | Western Pacific | Observational | 80 | 50 HT, 30 Control | 61.8 (10.6) |
| Nuli a<sup>14</sup> | China | UMICs | Western Pacific | Observational | 60 | 20 T2DM, 20 Impaired glucose regulation, 20 Control | NR |
| Nuli b<sup>15</sup> | China | UMICs | Western Pacific | Observational | 561 | 145 T2DM, 138 IFG, 278 Control | 47.08 (10.43) |
| Qiu<sup>16</sup> | China | UMICs | Western Pacific | Observational | 55 | 28 Obesity, 27 Control | 37.75 (12.54) |
| Tao<sup>17</sup> | China | UMICs | Western Pacific | Observational | 56 | 14 DN, 14 T2DM without DN, 28 Control | 50.84 (12.30) |
| Wang<sup>18</sup> | China | UMICs | Western Pacific | Observational | 46 | 26 Obesity 20 Healthy | 34.2 (10.1) |
| Wu<sup>19</sup> | China | UMICs | Western Pacific | Observational | 49 | 23 GDM, 26 Non-GDM | GDM Mean 36 (IQR 32-38.5), Non-GDM Mean 32.5 (IQR 30-35) |
| Xu<sup>20</sup> | China | UMICs | Western Pacific | Observational | 61 | 30 GDM, 31 Control | 33.0 (4.5) |
| Zhang<sup>21</sup> | China | UMICs | Western Pacific | Observational | 180 | 130 T2DM, 50 Healthy | 58.7 (9.4) |
| Zhao<sup>22</sup> | China | UMICs | Western Pacific | Observational | 58 | 25 Obesity with NAFLD, 18 Obesity only, 15 Healthy | 13.7 (1.8) |
| Zheng<sup>23</sup> | China | UMICs | Western Pacific | Observational | 99 | 36 DLP, 63 Healthy | 36 (NR) |
| Zhou<sup>52</sup> | China | UMICs | Western Pacific | Quasi-exp | 30 | 30 Obesity or Overweight | 47.9 (5.4) |
| Zuo b<sup>24</sup> | China | UMICs | Western Pacific | Observational | 49 | 34 HT, 15 Healthy | HT Med 54.5 (IQR 49.25, 57.75), Healthy Med 58 (IQR 52, 60) |
| Cuesta-Zuluaga<sup>25</sup> | Colombia | UMICs | Americas | Observational | 441 | 30% Obesity (132), 41% Central Obesity (180), 59% HT (259) | 41 (1) |
| Kern<sup>44</sup> | Denmark | HICs | European | RCT | 88 | 88 Overweight or Obesity | Med 36 (IQR 30, 41) |
| Salah<sup>26</sup> | Egypt | LMICs | Eastern Mediterranean | Observational | 60 | 25 Obesity with T2DM, 25 Obesity only, 5 T2DM only, 5 Control | 43.9 (13.3) |
| Olsson<sup>27</sup> | France | HICs | European | Observational | 58 | 34 Obesity, 24 non-Obesity | 42.3 (16.3) |
| Seck<sup>28</sup> | France | HICs | European | Observational | 56 | 17 Obesity, 39 Lean | NR |
| Frost<sup>55</sup> | Germany | HICs | European | Quasi-exp | 12 | 12 T2DM with Obesity | 57.2 (NR) |
| Kellerer<sup>53</sup> | Germany | HICs | European | Quasi-exp | 17 | 17 Morbid Obesity | 41.8 (9.1) |
| Ejtahed<sup>49</sup> | Iran | UMICs | Eastern Mediterranean | RCT | 36 | 36 Non-Diabetic Obese Woman | 36 (7) |
| Conterno<sup>42</sup> | Italy | HICs | European | RCT | 62 | 62 Hypercholesterolemia | 48.5 (9.0) |
| Ponzo<sup>29</sup> | Italy | HICs | European | Observational | 29 | 29 GDM Offspring | 3-5 Day of Life |
| Uemura<sup>46</sup> | Japan | HICs | Western Pacific | RCT | 44 | 44 Obesity | 62.7 (8.8) |
| Vera<sup>45</sup> | Mexico | UMICs | Americas | RCT | 81 | 81 T2DM | Range 30-60 |
| Bakker<sup>51</sup> | Netherlands | HICs | European | Quasi-exp | 20 | 10 Obesity with Metabolic Syndrome, 10 Lean | 43.6 (17.2) |
| vanBommel<sup>47</sup> | Netherlands | HICs | European | RCT | 44 | 44 T2DM | NR |
| Belkova<sup>30</sup> | Russia | UMICs | European | Observational | 30 | 12 Obesity, 18 Normal weight | 14.6 (1.6) |
| Koo<sup>31</sup> | Singapore | HICs | Western Pacific | Observational | 35 | 5 Metabolic Syndrome, 9 Obesity, 2 T2DM, 22 Central Obesity | Med 39 (Range 22-70) |
| Velikonja<sup>48</sup> | Slovenia | HICs | European | RCT | 43 | 43 Metabolic syndrome/High risk for Metabolic Syndrome | 52.07 (7.71) |
| Kim<sup>41</sup> | South Korea | HICs | Western Pacific | Observational | 766 | 313 NAFLD, 453 Control | 44.9 (8.1) |
| Romo-Vaquero<sup>54</sup> | Spain | HICs | European | Quasi-exp | 249 | 49 Obesity/Overweight, 200 Healthy | 42.6 (11.5) |
| Lin<sup>32</sup> | Taiwan | HICs | Western Pacific | Observational | 20 | 20 Obesity | 37.11 (10.0) |
| Astbury<sup>33</sup> | UK | HICs | European | Observational | 141 | 65 Biopsy-proven NASH, 76 Healthy | 64.19 (2.96) |
| Faucher<sup>34</sup> | USA | HICs | Americas | Observational | 21 | 15 Obesity, 6 Normal | 29.7 (5.1) |
| Fei<sup>35</sup> | USA | HICs | Americas | Observational | 655 | 229 Obesity, 149 Overweight, 277 Normal | 34.9 (6.4) |
| Karvonen<sup>50</sup> | USA | HICs | Americas | RCT | 502 | 149 Obesity, 353 Normal | 3 (NR) |
| Maskarinec<sup>37</sup> | USA | HICs | Americas | Observational | 1735 | 30.3% Obesity, 40.4% Overweight, 29.3% Normal | 69 (NR) |
| Schwimmer<sup>38</sup> | USA | HICs | Americas | Observational | 124 | 87 Biopsy-proven NAFLD, 37 Obesity without NAFLD | NAFLD Med 12 (IQR 10,14), Obesity without NAFLD Med 13 (IQR 11,14) |
| Shen<sup>39</sup> | USA | HICs | Americas | Observational | 26 | 26 Severe Obesity | NR |
| Stanislawski<sup>36</sup> | USA | HICs | Americas | Observational | 152 | 103 Obesity, 49 Control | 34.0 (12.2) |
| Sun<sup>40</sup> | USA | HICs | Americas | Observational | 529 | 35.1% HT | 55.3 (3.4) |

DLP, dyslipidemia; DN, diabetes nephropathy; GDM, gestational diabetes mellitus; HICs, high- income countries; HT, hypertension; IFG, impaired fasting glucose; IQR, interquartile range; LMICs, lower-middle-income countries; Med, median; NAFLD, nonalcoholic fatty liver disease; NR, not reported; Quasi-exp, quasi-experimental; RCT, randomized controlled trial; SD, standard deviation; T1DM, type I diabetes mellitus; T2DM, type II diabetes mellitus; UMICs, upper-middle- income countries; WHO, World Health Organization.

According to WHO region, there were 21 studies conducted in Western Pacific Region,^8 9 11-24 31 32 41 46 52^ 14 studies in European Region,^27-30 33 42-44 47 48 51 53-55^ 10 studies in Region of the Americas,^25 34-40 45 50^ and two studies conducted in Eastern Mediterranean Region.^26 49^ According to the World Bank,^56^ there were 27 studies conducted in high-income countries,^9 11 27-36 38-44 47 48 50 51 53-55 57^ 19 in upper-middle income countries,^10 12-25 30 45 49 52^ and one in low-middle income countries.^26^

### Gut microbiome diversity measures

Of 47 included studies, there were 35 studies reported both alpha and beta diversities, 12 studies reported alpha diversity only, and no study reported beta diversity only. Of 13 alpha diversity measures, the Shannon index was the most commonly used in 37 studies (78.7%),^10-24 26 28-36 38 40 41 44-48 50 52 53 57^ followed by Chao1 index (19 studies),^10-13 15-18 23 24 27 29 30 32 35 42 43 48 52^ Operational Taxonomic Unit (OTU) richness (15 studies),^9 10 12 13 17 25-28 34 35 49 50 52 53^ Simpson index (13 studies),^12 13 15-17 20 23 30 31 46 49 52 55^ Abundance-based Coverage Estimators (ACE) index (10 studies),^12 13 15 17 18 20 30 32 52 54^ Observed species (9 studies),^13-15 17 18 20 29 36 52^ Faith’s Phylogenetic diversity (7 studies),^18 36 39 41 47 51 52^ Phylogenetic diversity (5 studies),^10 16 27 55 58^ Good’s coverage (3 studies),^12 13 18^ Pielou’s evenness index (2 studies),^24 27^ Amplicon Sequence Variant (ASV) richness (2 studies),^41 44^ inversed Simpson index (2 studies),^9 34^ and Fisher alpha index (1 study).^34^

Of 2 beta diversity measures, the UniFrac was the most commonly used in 24 studies including unweighted UniFrac (17 studies),^12 14 18 20 21 25-27 29 33-35 39 41 45 52 57^ and weighted UniFrac (16 studies),^12 13 16-18 20 25 26 34 35 37 41 44 48 51 53^ followed by Bray-Curtis dissimilarity (16 studies).^11 16 17 19 20 23 31 38 40-44 47 49 55^ The summary result was shown in table 2 and result for each included study was provided in online supplemental appendix 2.

**Table 2.**
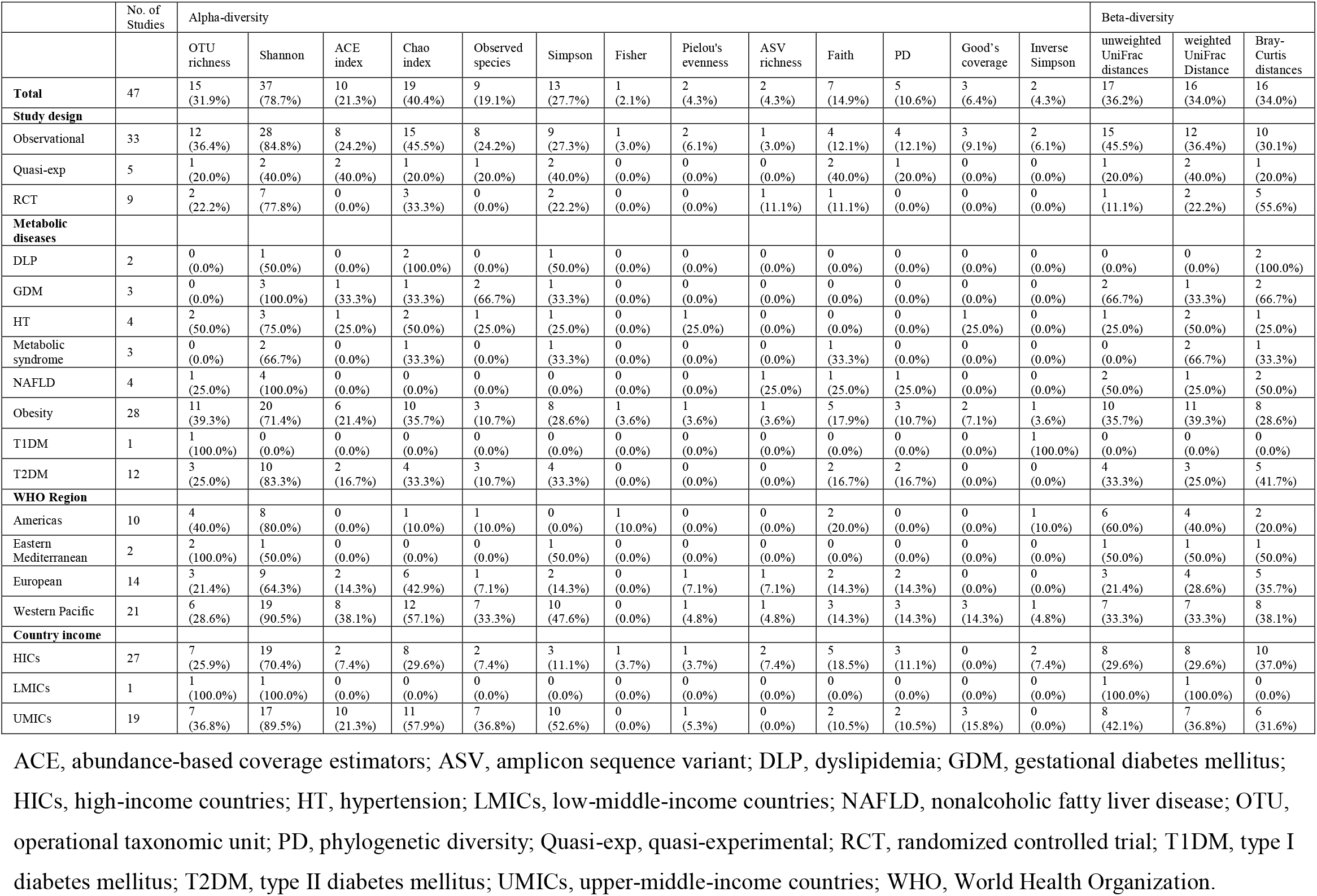
Gut microbiome diversity index usage in metabolic disease articles

|  | No. of Studies | Alpha-diversity |  |  |  |  |  |  |  |  |  |  |  |  | Beta-diversity |  |  |
| --- | --- | --- | --- | --- | --- | --- | --- | --- | --- | --- | --- | --- | --- | --- | --- | --- | --- |
|  |  | OTU richness | Shannon | ACE index | Chao index | Observed species | Simpson | Fisher | Pielou's evenness | ASV richness | Faith | PD | Good's coverage | Inverse Simpson | unweighted UniFrac distances | weighted UniFrac Distance | Bray-Curtis distances |
| **Total** | 47 | 15 (31.9%) | 37 (78.7%) | 10 (21.3%) | 19 (40.4%) | 9 (19.1%) | 13 (27.7%) | 1 (2.1%) | 2 (4.3%) | 2 (4.3%) | 7 (14.9%) | 5 (10.6%) | 3 (6.4%) | 2 (4.3%) | 17 (36.2%) | 16 (34.0%) | 16 (34.0%) |
| **Study design** |  |  |  |  |  |  |  |  |  |  |  |  |  |  |  |  |  |
| Observational | 33 | 12 (36.4%) | 28 (84.8%) | 8 (24.2%) | 15 (45.5%) | 8 (24.2%) | 9 (27.3%) | 1 (3.0%) | 2 (6.1%) | 1 (3.0%) | 4 (12.1%) | 4 (12.1%) | 3 (9.1%) | 2 (6.1%) | 15 (45.5%) | 12 (36.4%) | 10 (30.1%) |
| Quasi-exp | 5 | 1 (20.0%) | 2 (40.0%) | 2 (40.0%) | 1 (20.0%) | 1 (20.0%) | 2 (40.0%) | 0 (0.0%) | 0 (0.0%) | 0 (0.0%) | 2 (40.0%) | 1 (20.0%) | 0 (0.0%) | 0 (0.0%) | 1 (20.0%) | 2 (40.0%) | 1 (20.0%) |
| RCT | 9 | 2 (22.2%) | 7 (77.8%) | 0 (0.0%) | 3 (33.3%) | 0 (0.0%) | 2 (22.2%) | 0 (0.0%) | 0 (0.0%) | 1 (11.1%) | 1 (11.1%) | 0 (0.0%) | 0 (0.0%) | 0 (0.0%) | 1 (11.1%) | 2 (22.2%) | 5 (55.6%) |
| **Metabolic diseases** |  |  |  |  |  |  |  |  |  |  |  |  |  |  |  |  |  |
| DLP | 2 | 0 (0.0%) | 1 (50.0%) | 0 (0.0%) | 2 (100.0%) | 0 (0.0%) | 1 (50.0%) | 0 (0.0%) | 0 (0.0%) | 0 (0.0%) | 0 (0.0%) | 0 (0.0%) | 0 (0.0%) | 0 (0.0%) | 0 (0.0%) | 0 (0.0%) | 2 (100.0%) |
| GDM | 3 | 0 (0.0%) | 3 (100.0%) | 1 (33.3%) | 1 (33.3%) | 2 (66.7%) | 1 (33.3%) | 0 (0.0%) | 0 (0.0%) | 0 (0.0%) | 0 (0.0%) | 0 (0.0%) | 0 (0.0%) | 0 (0.0%) | 2 (66.7%) | 1 (33.3%) | 2 (66.7%) |
| HT | 4 | 2 (50.0%) | 3 (75.0%) | 1 (25.0%) | 2 (50.0%) | 1 (25.0%) | 1 (25.0%) | 0 (0.0%) | 1 (25.0%) | 0 (0.0%) | 0 (0.0%) | 0 (0.0%) | 1 (25.0%) | 0 (0.0%) | 1 (25.0%) | 2 (50.0%) | 1 (25.0%) |
| Metabolic syndrome | 3 | 0 (0.0%) | 2 (66.7%) | 0 (0.0%) | 1 (33.3%) | 0 (0.0%) | 1 (33.3%) | 0 (0.0%) | 0 (0.0%) | 0 (0.0%) | 1 (33.3%) | 0 (0.0%) | 0 (0.0%) | 0 (0.0%) | 0 (0.0%) | 2 (66.7%) | 1 (33.3%) |
| NAFLD | 4 | 1 (25.0%) | 4 (100.0%) | 0 (0.0%) | 0 (0.0%) | 0 (0.0%) | 0 (0.0%) | 0 (0.0%) | 0 (0.0%) | 1 (25.0%) | 1 (25.0%) | 1 (25.0%) | 0 (0.0%) | 0 (0.0%) | 2 (50.0%) | 1 (25.0%) | 2 (50.0%) |
| Obesity | 28 | 11 (39.3%) | 20 (71.4%) | 6 (21.4%) | 10 (35.7%) | 3 (10.7%) | 8 (28.6%) | 1 (3.6%) | 1 (3.6%) | 1 (3.6%) | 5 (17.9%) | 3 (10.7%) | 2 (7.1%) | 1 (3.6%) | 10 (35.7%) | 11 (39.3%) | 8 (28.6%) |
| T1DM | 1 | 1 (100.0%) | 0 (0.0%) | 0 (0.0%) | 0 (0.0%) | 0 (0.0%) | 0 (0.0%) | 0 (0.0%) | 0 (0.0%) | 0 (0.0%) | 0 (0.0%) | 0 (0.0%) | 0 (0.0%) | 1 (100.0%) | 0 (0.0%) | 0 (0.0%) | 0 (0.0%) |
| T2DM | 12 | 3 (25.0%) | 10 (83.3%) | 2 (16.7%) | 4 (33.3%) | 3 (10.7%) | 4 (33.3%) | 0 (0.0%) | 0 (0.0%) | 0 (0.0%) | 2 (16.7%) | 2 (16.7%) | 0 (0.0%) | 0 (0.0%) | 4 (33.3%) | 3 (25.0%) | 5 (41.7%) |
| **WHO Region** |  |  |  |  |  |  |  |  |  |  |  |  |  |  |  |  |  |
| Americas | 10 | 4 (40.0%) | 8 (80.0%) | 0 (0.0%) | 1 (10.0%) | 1 (10.0%) | 0 (0.0%) | 1 (10.0%) | 0 (0.0%) | 0 (0.0%) | 2 (20.0%) | 0 (0.0%) | 0 (0.0%) | 1 (10.0%) | 6 (60.0%) | 4 (40.0%) | 2 (20.0%) |
| Eastern Mediterranean | 2 | 2 (100.0%) | 1 (50.0%) | 0 (0.0%) | 0 (0.0%) | 0 (0.0%) | 1 (50.0%) | 0 (0.0%) | 0 (0.0%) | 0 (0.0%) | 0 (0.0%) | 0 (0.0%) | 0 (0.0%) | 0 (0.0%) | 1 (50.0%) | 1 (50.0%) | 1 (50.0%) |
| European | 14 | 3 (21.4%) | 9 (64.3%) | 2 (14.3%) | 6 (42.9%) | 1 (7.1%) | 2 (14.3%) | 0 (0.0%) | 1 (7.1%) | 1 (7.1%) | 2 (14.3%) | 2 (14.3%) | 0 (0.0%) | 0 (0.0%) | 3 (21.4%) | 4 (28.6%) | 5 (35.7%) |
| Western Pacific | 21 | 6 (28.6%) | 19 (90.5%) | 8 (38.1%) | 12 (57.1%) | 7 (33.3%) | 10 (47.6%) | 0 (0.0%) | 1 (4.8%) | 1 (4.8%) | 3 (14.3%) | 3 (14.3%) | 3 (14.3%) | 1 (4.8%) | 7 (33.3%) | 7 (33.3%) | 8 (38.1%) |
| **Country income** |  |  |  |  |  |  |  |  |  |  |  |  |  |  |  |  |  |
| HICs | 27 | 7 (25.9%) | 19 (70.4%) | 2 (7.4%) | 8 (29.6%) | 2 (7.4%) | 3 (11.1%) | 1 (3.7%) | 1 (3.7%) | 2 (7.4%) | 5 (18.5%) | 3 (11.1%) | 0 (0.0%) | 2 (7.4%) | 8 (29.6%) | 8 (29.6%) | 10 (37.0%) |
| LMICs | 1 | 1 (100.0%) | 1 (100.0%) | 0 (0.0%) | 0 (0.0%) | 0 (0.0%) | 0 (0.0%) | 0 (0.0%) | 0 (0.0%) | 0 (0.0%) | 0 (0.0%) | 0 (0.0%) | 0 (0.0%) | 0 (0.0%) | 1 (100.0%) | 1 (100.0%) | 0 (0.0%) |
| UMICs | 19 | 7 (36.8%) | 17 (89.5%) | 10 (21.3%) | 11 (57.9%) | 7 (36.8%) | 10 (52.6%) | 0 (0.0%) | 1 (5.3%) | 0 (0.0%) | 2 (10.5%) | 2 (10.5%) | 3 (15.8%) | 0 (0.0%) | 8 (42.1%) | 7 (36.8%) | 6 (31.6%) |

ACE, abundance-based coverage estimators; ASV, amplicon sequence variant; DLP, dyslipidemia; GDM, gestational diabetes mellitus; HICs, high-income countries; HT, hypertension; LMICs, low-middle-income countries; NAFLD, nonalcoholic fatty liver disease; OTU, operational taxonomic unit; PD, phylogenetic diversity; Quasi-exp, quasi-experimental; RCT, randomized controlled trial; T1DM, type I diabetes mellitus; T2DM, type II diabetes mellitus; UMICs, upper-middle-income countries; WHO, World Health Organization.

### Study design and gut microbiome diversity measures

Most common microbiome alpha-diversity measures for observational studies and randomized controlled trials were Shannon diversity in 28 observational studies (84.8%),^10-24 26 28-31 33-38 40 41^ and 7 RCTs (77.8%) respectively ^43-48 50^ Most common microbiome alpha-diversity measures for quasi-experimental were Shannon diversity,^52 53^ ACE index,^52 54^ Simpson index,^52 55^ and Faith’s Phylogenetic diversity in 2 studies (40%).^51 52^ Good’s coverage (3 studies),^12 13 18^ and Pielou’s evenness index (2 studies) were reported only in observational studies.^27 52^

Most common microbiome beta-diversity measures were different among study designs. For observational studies, unweighted Unifrac was most common with reported in 45.5% of included observational studies,^12 14 18 20 21 25-27 29 33-35 37 39 41^ Bray-Curtis dissimilarity for randomized controlled trials (55.6%),^42-44 48 49^ and weighted Unifrac for quasi-experimental studies (40%).^51 53^

### Type of metabolic diseases and gut microbiome diversity measures

Most common microbiome alpha-diversity for all metabolic diseases was Shannon diversity. Most common microbiome beta-diversity measures for obesity and type II diabetes mellitus were weighted Unifrac (39.3%),^12 16 17 25 26 34 35 44 51 53 57^ and Bray-Curtis dissimilarity (33.3%) respectively.^17 43 47 55^

### Geographical location and gut microbiome diversity measures

Most common microbiome alpha-diversity for studies in Western Pacific, European, and America Regions was Shannon diversity. Most common microbiome alpha-diversity for Eastern Mediterranean Region was OTU richness. Good’s coverage (3 studies) were used only in Western Pacific Region.^12 13 18^

Most common microbiome beta-diversity measures for Western Pacific, European and America were Bray-Curtis dissimilarity (38.1%),^11 16 17 19 20 23 31 41^ Bray-Curtis dissimilarity (35.7%),^42-44 47 55^ and unweighted Unifrac (60%) respectively.^25 34 35 37 39 45^ For microbiome beta-diversity measures for Eastern Mediterranean, all beta-diversity were equal in number (1 study, 50%).

### Country income and gut microbiome diversity measures

Most common microbiome alpha-diversity for all type of country incomes was Shannon diversity. Amplicon Sequence Variant (ASV) richness (2 studies),^41 44^ and inversed Simpson index (2 studies) were both used in only HICs.^29 36^ Good’s coverage (3 studies) were used only in UMICs.^12 13 18^ There was a variation of most common microbiome beta-diversity measures among different country incomes. For HICs, Bray-Curtis dissimilarity was the most common beta diversity measure (37%),^11 31 38 40-44 47 55^ unweighted Unifrac for UMICs (42.1%),^12 14 18 20-22 25 45 52^ and both weighted and unweighted Unifrac for LMIC (100%).^26^

## DISCUSSION

To our knowledge, this is the first systematic review evaluating the choice of gut microbiome diversity measurements in patients with metabolic conditions. The systematic review process identified 47 articles that met the inclusion and exclusion criteria. A meta-analysis was not performed due to the purpose of this study to identify the usage of gut microbiome diversity measurement in metabolic diseases studies. The result suggested that there were variations in measures of gut microbiome diversity in the metabolic disease literatures. For alpha-diversity, there were 13 different measurement methods used for the analysis of gut microbiota. The Shannon index was the most commonly used which presented in 37 studies (78.7%) while other methods including Chao1 index, Operational Taxonomic Unit (OTU) richness, Simpson index, Faith’s Phylogenetic diversity, and Abundance-based Coverage Estimators (ACE) index were used only 20-40%. For beta-diversity, the UniFrac was the most commonly used which assessed in 24 studies (unweighted 17 studies and weighted 16 studies), followed by Bray-Curtis dissimilarity (16 studies). All beta-diversity measures were used in a similar quantity.

Alpha diversity measures the diversity within a sample diversity and is based on the relative abundance of taxa; for example, species or OTUs.^59^ Alpha diversity are used to identify the richness (number of taxonomic groups), evenness (distribution of abundances of the groups), or both. There are three subtypes of alpha diversity by purpose of estimation including richness estimators, richness and evenness estimators, and phylogenetic richness estimators. The difference between these estimators was shown in online supplemental table 1. Species richness refers to the number of different species present in a community while evenness compares the uniformity of the population of the species (figure 2).

**Figure 2.**
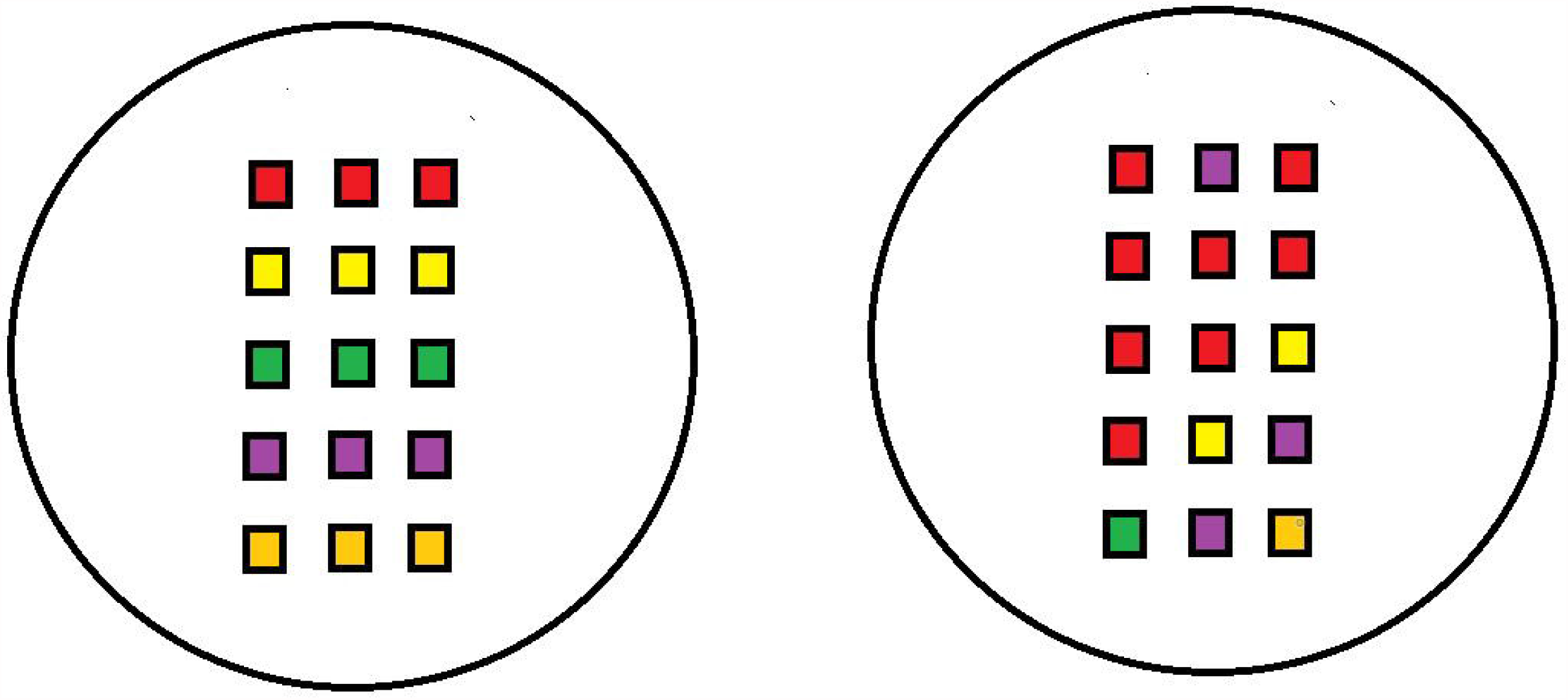
Community A and community B have the same species richness, five species each. The organisms in community A are more evenly distributed than in community B

Of all included studies, Shannon diversity was the most common alpha diversity measurement. Shannon diversity is an example of the richness and evenness estimators which the value of diversity increases both when the number of species increases and when evenness increases. It is a measurement of entropy and the uncertainty of the sampling outcome. Shannon diversity also contemplates the relative abundances of different species.^60^ The advantages of Shannon diversity are simplicity and appropriateness for the community dominant by two or three species. However, Shannon diversity weighs more on species richness which causes measure of the character of the species abundance distribution (evenness) less sensitive.

Another richness and evenness estimator is Simpson’s diversity. The advantage of Simpson’s diversity is simplicity while the drawback is insensitivity of the species richness measurement. Due to simplicity and ability to measure both richness and evenness, Shannon diversity and Simpson’s diversity are widely used in all studies.^58^

Another type of the estimators is the richness estimator which includes OTU richness, Chao1 index, and ACE index. OTU richness is defined as the count of different species represented in a community. Similar to Shannon and Simpson’s diversity, the advantage of OTU richness is simplicity which makes OTU richness in top three of alpha diversity measurement usage. However, the disadvantage of OTU richness is sensitivity to sample size. In our studies, Chao1 index was used in 19 studies (40.4%) and ACE index in 10 studies (21.3%). Richness estimators evaluate the total richness of a community.^61^ Chao1 and ACE have been developed to estimate richness from abundance data. They are the indicators of species richness that is sensitive to rare OTUs. Chao1 is based on the theory that rare species provide the most information about the number of missing species which is useful for rare species and performs accurately if the sample size is reasonably large. Therefore, Chao1 index is particularly useful for the low-abundance species while the drawback is underestimation of rich and highly heterogeneous species.

Finally, phylogenetic diversity (PD) is defined as the connecting of all organisms in a phylogenetic tree which estimates diversity across a tree and provides a phylogenetic analog of taxonomic diversity.^62^ PD provides a convenient, evolutionary measure of diversity that does not depend on the ability to identify species count which ultimately leads to a complex and relatively stable community of microorganisms.^63^ However, the limitation is lack of sequence data for the large majority of species which requires a molecular laboratory with proper facilities, thus the cost is inevitably highed.^64^

Beta-diversity is the measure for differences between samples from different groups. The overall community composition and structure are identified by this measure. Beta-diversity in our research including Unifrac distance and Bray-Curtis dissimilarity are similar in usage quantities. “UniFrac” considers the phylogenetic relationships between the microbes found in two samples (Similar to phylogenetic diversity) which provides a convenient measure of diversity that does not depend on the ability to identify species count.^65^ It estimates differences between samples or groups based on phylogenetic distance. Unifrac distance is divided into unweighted and weighted Unifrac. Unweighted UniFrac is the fraction of branch lengths between all microbes in both samples that are different between the samples.^65 66^ Weighted UniFrac is similar to unweighted UniFrac but takes the abundances of microbes in the samples into account. Weighted UniFrac is largely impacted by the abundances of the microbes while unweighted UniFrac does not take abundance into account. Both unweighted Unifrac and weighted Unifrac are used to determine whether communities were significantly different. Unweighted Unifrac is sensitive to detect microbial richness changes in rare species while weighted Unifrac can incorporate the abundance information and reduce the rare species’ contribution. However, both measures should not be used as a distance metric for multivariate statistical analyses.^66 67^

Bray-Curtis dissimilarity shows the microbes’ abundances which are shared between two samples. Bray-Curtis dissimilarity quantifies the dissimilarity between two samples or groups ranged from 0 to 1 which is not a true distance. For example, if both samples have the same number of microbes at the same abundance, their dissimilarity will equal zero. On the other hand, the dissimilarity will equal 1 if two samples have definitely no shared microbes. The benefit of Bray-Curtis dissimilarity is that it gives more weight to common species. Moreover, Bray-Curtis dissimilarity is simple and doesn’t make assumptions about genetic relationships.^68^

There were several limitations in this systematic scoping review. First, this systematic scoping review aims to provide evidence on the usage of gut microbiome diversity measurement in metabolic diseases studies, thus meta-analysis is not planned to perform. Second, this study focused on the gut microbiome diversity measures in patients with metabolic conditions only. The generalizability should be considered when applying the results in studies on other diseases.

In conclusion, this systematic scoping review and meta-analysis provided the first evidence on the gut microbiome Diversity Measures for Metabolic Conditions. Alpha-diversity and beta-diversity are innovative tools for gut microbiota analysis. Various measurement of gut microbiome diversity have been used in the literature. All measurements have unique characteristics, advantages, and disadvantages which lead to different usage frequency. The measures were chosen considering cost, simplicity, and types of research.

## Data Availability

All data relevant to the study are included in the article or uploaded as supplementary information.

## Acknowledgement

The authors are thankful to the Prince Mahidol Award Youth Program and Professor Nijasri C. Suwanwela for their research and academic assistance, as well as Miss Pim Sermsaksasithorn for providing mental support.

## Contributors

CS, and KP conceived and designed the research. CS and TN performed data acquisition, analyzed and interpretated data, and drafted the manuscript. KP made critical revisions related to important intellectual content of the manuscript. All authors have read and approved the final version of manuscript.

## Patient consent for publication

Not required.

## Ethics approval

The study required no Ethics Committee approval as long it is a systematic review study and no human subject was directly involved.

## Supplemental material

Supplemental appendix 1. Full search strategy

Supplemental appendix 2. Measurement methods for each included study

Supplemental table 1. Estimator characteristics

