## Supplemental appendix 1 for "Gut microbiome diversity measures for metabolic conditions: a systematic scoping review"

**Supplementary File 1. Full search strategy**

| Set # | PubMed | Results |
| --- | --- | --- |
| 1  Gut Microbiome | "gastrointestinal microbiome"[MeSH Terms] OR ((gut[tiab] OR intestin*[tiab] OR gastrointestin*[tiab] OR colon*[tiab] OR rectal[tiab] OR rectum[tiab] OR stool[tiab] OR feces[tiab] OR faeces[tiab] OR fecal[tiab] OR faecal[tiab]) AND ("microbiota"[MeSH Terms] OR "mycobiome"[MeSH Terms] OR microbiome*[tiab] OR microbiota*[tiab] OR microbial[tiab] OR microbe*[tiab] OR microflora*[tiab] OR flora*[tiab] OR microorganism*[tiab] OR pathobiont*[tiab] OR mycobiome*[tiab] OR mycobiota*[tiab] OR virome*[tiab] OR phylotype*[tiab])) OR enterotype*[tiab] | **89194** |
| 2  Diversity | diversit*[tiab] OR abundan*[tiab] OR richness*[tiab] OR evenness*[tiab] OR dissimilarit*[tiab] | **489439** |
| 3 | #1 AND #2 | **16344** |
| 4 | animals[MeSH Terms] NOT humans[MeSH Terms] | **4682770** |
| 5 | #3 NOT #4 | **11807** |
| 6 | English[lang] | **26086457** |
| 7 | #5 AND #6 | **11634** |
| 8 | "2019/01/01"[PDAT] : "2019/12/31"[PDAT] | **1402266** |
| 9 | #7 AND #8 | **3111** |

| Set # | Embase | Results |
| --- | --- | --- |
| 1  Gut Microbiome | 'intestine flora'/exp OR (('gut':ti,ab OR 'intestin*':ti,ab OR 'gastrointestin*':ti,ab OR 'colon*':ti,ab OR 'rectal':ti,ab OR 'rectum':ti,ab OR 'stool':ti,ab OR 'feces':ti,ab OR 'faeces':ti,ab OR 'fecal':ti,ab OR 'faecal':ti,ab) AND ('microbiome'/exp OR 'microflora'/exp OR 'mycobiome'/exp OR 'microbiome*':ti,ab OR 'microbiota*':ti,ab OR 'microbial':ti,ab OR 'microbe*':ti,ab OR 'microflora*':ti,ab OR 'flora*':ti,ab OR 'microorganism*':ti,ab OR 'pathobiont*':ti,ab OR 'mycobiome*':ti,ab OR 'mycobiota*':ti,ab OR 'virome*':ti,ab OR 'phylotype*':ti,ab)) OR 'enterotype*':ti,ab | **126127** |
| 2  Diversity | 'diversit*':ti,ab OR 'abundan*':ti,ab OR 'richness*':ti,ab OR 'evenness*':ti,ab OR 'dissimilarit*':ti,ab | **549572** |
| 3 | #1 AND #2 | **21632** |
| 4 | [animals]/lim NOT [humans]/lim | **5765837** |
| 5 | #3 NOT #4 | **13783** |
| 6 | english:la | **31021053** |
| 7 | #5 AND #6 | **13542** |
| 8 | [2019]/py | **1585050** |
| 9 | #7 AND #8 | **2536** |

| Set # | CENTRAL | Results |
| --- | --- | --- |
| 1  Gut Microbiome | [mh "gastrointestinal microbiome"] OR ((gut:ti,ab,kw OR intestin*:ti,ab,kw OR gastrointestin*:ti,ab,kw OR colon*:ti,ab,kw OR rectal:ti,ab,kw OR rectum:ti,ab,kw OR stool:ti,ab,kw OR feces:ti,ab,kw OR faeces:ti,ab,kw OR fecal:ti,ab,kw OR faecal:ti,ab,kw) AND ([mh microbiota] OR [mh mycobiome] OR microbiome*:ti,ab,kw OR microbiota*:ti,ab,kw OR microbial:ti,ab,kw OR microbe*:ti,ab,kw OR microflora*:ti,ab,kw OR flora*:ti,ab,kw OR microorganism*:ti,ab,kw OR pathobiont*:ti,ab,kw OR mycobiome*:ti,ab,kw OR mycobiota*:ti,ab,kw OR virome*:ti,ab,kw OR phylotype*:ti,ab,kw)) OR enterotype*:ti,ab,kw | **8915** |
| 2  Diversity | diversit*:ti,ab,kw OR abundan*:ti,ab,kw OR richness*:ti,ab,kw OR evenness*:ti,ab,kw OR dissimilarit*:ti,ab,kw | **4452** |
| 3 | #1 AND #2 | **1067** |
| 4 | [mh animals] NOT [mh humans] | **6950** |
| 5 | #3 NOT #4 | **1051** |
| 6 | #5 restrict publication year to 2019 | **282** |
